# Venous Access Problems in Sickle Cell Disease Patients: Results of Upper Extremity Thrombolysis in the setting of Catheter Related Thrombosis

**DOI:** 10.1101/2020.05.31.20117622

**Authors:** Jonathan G. Martin

## Abstract

**Purpose:** Sickle cell disease is the most common monogenic disorder. All the elements of Virchow’s triad – hypercoagulability, endothelial dysfunction and impaired blood flow – are present in sickle cell disease patients, and can lead to thrombosis and vaso-occlusive crises. Central venous catheters are commonly used in sickle cell disease patients for rapid transfusion to avoid a vaso-occlusive crisis or for treatment after crises onset. However, central venous catheters themselves are an additional source for thrombus formation. We investigate a single day thrombolysis protocol for the treatment of catheter related thrombosis.

**Materials and Methods:** We present an Institutional Review Board approved retrospective analysis spanning from June 2016 to April 2018 evaluating upper extremity thrombolysis in the setting of catheter related thrombosis. All patients underwent a similar protocol involving peripheral upper extremity access, pharmomechanical thrombolysis, and angioplasty, which is described in detail. In 33% of the procedures, a recalcitrant stenosis was then stented. Maximal balloon size and stent sizes are included for reference.

**Results:** In follow up, one patient had a severe complication possibly related to the large thrombus burden, and expired. One patient who did not initially have recalcitrant stenosis and therefor did not have a stent placed during the initial procedure, had recurrent stenosis and thrombosis 6 months after the initial procedure, and had repeat pharmomechanical thrombolysis performed, with stent placement at that time. The other procedures were without complication, and the upper extremity and central venous systems remained patent at most recent follow-up, up to 30 months post procedure.

**Conclusions:** In the circumstance of severe catheter related thrombosis, we present a protocol which can be considered for therapeutic use without further intervention required in 78% of cases at 2 years.

## Background

Sickle cell disease (SCD) is the most common monogenic disorder.^1^ SCD is most prevalent among sub-Saharan Africans (up to 25% gene frequency in some populations), and is also common in persons of Mediterranean, Saudi Arabian and Indian decent. In the United States, SCD affects approximately 100,000 persons and occurs in 1 of 365 Black or African-American births.^2^ Persons with SCD have a shortened life expectancy, approximately 42 years for females and 38 years for males.^3^ Despite this shortened life span, a combination of factors related to sickle cell disease and it treatments lead to a number of venous complications in sickle cell disease patients.

The genetic mutation which leads to sickle cell disease produces a substitution of valine for glutamic acid at the sixth codon of the β-globin chain.^4^ Homozygotes have sickle cell anemia (HbSS) and heterozygotes have sickle cell trait (HbAS).^5^ This mutation results in the production of HbS instead of HbA. In the deoxygenated condition, the molecules of hemoglobin S polymerize to form pseudo-crystalline structures known as ‘tactoids’.^6^ These crystalline RBCs are fragile and also inflexible. This inflexibility prevents their passage through the microcirculation. This leads to microvascular occlusion, which can lead to vaso-occlusive crisis, splenic infarction, acute chest syndrome, cerebrovascular complications, avascular necrosis, and priapism.^7^

Additional complications of sickle cell disease are directly or indirectly related to venous thromboembolism. All the elements of Virchow’s triad – hypercoagulability, endothelial dysfunction and impaired blood flow – are present in sickle cell disease patients.^8^ In SCD, excess hemoglobin is released in the circulation due to hemolysis. This free hemoglobin then reacts with nitric oxide to form inert nitrate, and erythrocyte arginase metabolizes arginine, the substrate for nitric oxide synthesis.^9^ Abnormal nitric oxide physiology and expression of various adhesive molecules on the surface of platelets eventually cause thromboembolism. In addition, several studies have identified that SCD patients have a higher circulating level of antiphospholipid antibodies and low levels of protein C and protein S, which further contribute to hypercoagulability^10^.

The largest thromboembolism study to date on SCD patients analyzed the National Hospital Discharge Survey and revealed that patients (< 40 years of age) with SCD have 3.5 times more probability of developing pulmonary embolism (PE) than their control groups^11^. Somewhat surprisingly, this same study showed that the prevalence of deep venous thrombosis (DVT) was not statistically different between SCD patients and controls. This may indicate that the higher probability of developing PE in SCD patients may be due to some local pulmonary vascular pathology.

Central venous catheters (CVCs) are commonly used in SCD patients for rapid transfusion to avoid a vaso-occlusive crisis or for treatment after crises onset. Although most commonly placed in the upper extremities, some SCD patients prefer placement in the femoral vein.^12^ CVCs are an additional source for thrombus formation, with an analysis of 25 prior studies demonstrating asymptomatic deep venous thrombosis (DVT) in as many as 41% of patients when venography was used to screen patients and 19% with screening ultrasound.^13^ Additionally, in a study using contrast venography in 114 patients 1 week after placement of a CVC, a DVT was detected in 53%, although this was only fully occlusive in 3%.^14^ Although the majority of asymptomatic catheter related thrombosis cases remain subclinical, symptomatic DVT occurs in 1%-5% of patients with a CVC^15^. Blockage or stenosis of catheters may also increase the chances of infection. In our patient population, we have seen at least one patient with septic emboli with their CVC as the source.

Prior reports of treatment for catheter related thrombosis are limited. Individual case studies have demonstrated successful thromboloysis, however these reports have limited follow up^16,17^. We set out to evaluate the outcomes of our one-day upper extremity thrombolysis protocol.

## Methods

This Health Insurance Portability and Accountability Act compliant retrospective study received Institutional Review Board approval and waiver of consent and included all patients who underwent upper extremity thrombolysis for catheter related thrombosis by Interventional Radiology (IR) at a single tertiary care medical center from June 2016 to April 2018. Information regarding patient demographics, past medical history, procedure indication, medications, labs, procedural details, and complications was acquired from the PACS and electronic medical record, primarily from IR procedural reports, IR or subspecialty clinic notes, inpatient admission and progress notes, consult notes, and lab results.

The primary outcome was recurrent thrombosis requiring re-intervention. Secondary outcomes were 30-day mortality, mortality related to the procedure, dissection and active extravasation, bleeding complications at the access site, infection at the access site, sepsis, and technical failure.

## Results

All patients underwent a similar one-day treatment protocol. Procedure planning was based on pre-procedure computed tomography of the chest with contrast (Figure 1). The brachial vein ipsilateral to the thrombus was accessed under ultrasound guidance. The access was upsized in standard fashion, and a 10 French sheath was placed. A 5 French diagnostic catheter was advanced centrally and, starting in the right atrium, a pullback venogram to the access site was performed (Figure 2). A multi side hole infusion catheter was then placed throughout the thrombus and 10 mg of TPA was slowly administered in to the clot, most commonly over a 30-minute duration. After completion of the TPA infusion, a CAT-8 Penumbra mechanical thrombectomy device was advanced and used to remove as much thrombus as possible, up to a total blood volume loss of 600 ml. Next, angiography was performed, and any areas of stenosis were treated with balloon angioplasty at the proceduralist’s discretion. All procedures included balloon angioplasty, with maximum balloon diameter included in Table 2. Patients with recalcitrant stenosis then underwent self-expanding stent placement, which was performed in 3 of the 9 procedures. All 3 stent placements were performed after the right femoral vein was accessed and “body floss” was obtained with a single wire coursing from the femoral vein access to the brachial vein access. If needed, the central venous catheter was snared internally and removed from the stent landing zone prior to stent deployment. The central venous catheter was again snared internally after stent deployment and repositioned through the inner channel of the stent within the superior cavo-atrial junction. Post procedure venography was performed to confirm patency (Figure 3).

**Table 1:** Summary of Patient Characteristics at time of procedure (9 procedures)

|  |  |
| --- | --- |
| Age | 39.3 +/- 15.3 yrs |
| Male | 7 (77.8%) |
| Female | 2 (22.2%) |
| African American or Black | 7 (77.8%) |
| Caucasian or White | 2 (22.2%) |
| Ipsilateral Port Catheter | 5 (55.6%) |
| Contralateral Port Catheter | 2 (22.2%) |
| Prior Catheter, No Current Catheter | 2 (22.2%) |

**Table 2:** Procedural details with maximum balloon diameter and uncovered stent size if required

| Procedure | Thrombosis Side | Port Catheter Side | Maximum balloon diameter | Resistant Stenosis with Stent Placement |
| --- | --- | --- | --- | --- |
| Procedure 1 | Right | Contralateral | 12 mm | No |
| Procedure 2 | Right | Ipsilateral | 12 mm | No |
| Procedure 3 | Left | None | 12 mm | No |
| Procedure 4 | Left | Ipsilateral | 10 mm | No |
| Procedure 5 | Left | Ipsilateral | 14 mm | No |
| Procedure 6 | Left | Contralateral | 12 mm | 14 x 40 mm |
| Procedure 7 | Right | None | 14 mm | No |
| Procedure 8 | Left | Ipsilateral | 14 mm | 14 x 80 mm |
| Procedure 9 | Left | Ipsilateral | 10 mm | Overlapping 14 x 80 mm and 14 x 40 mm |

**Figure 1:**
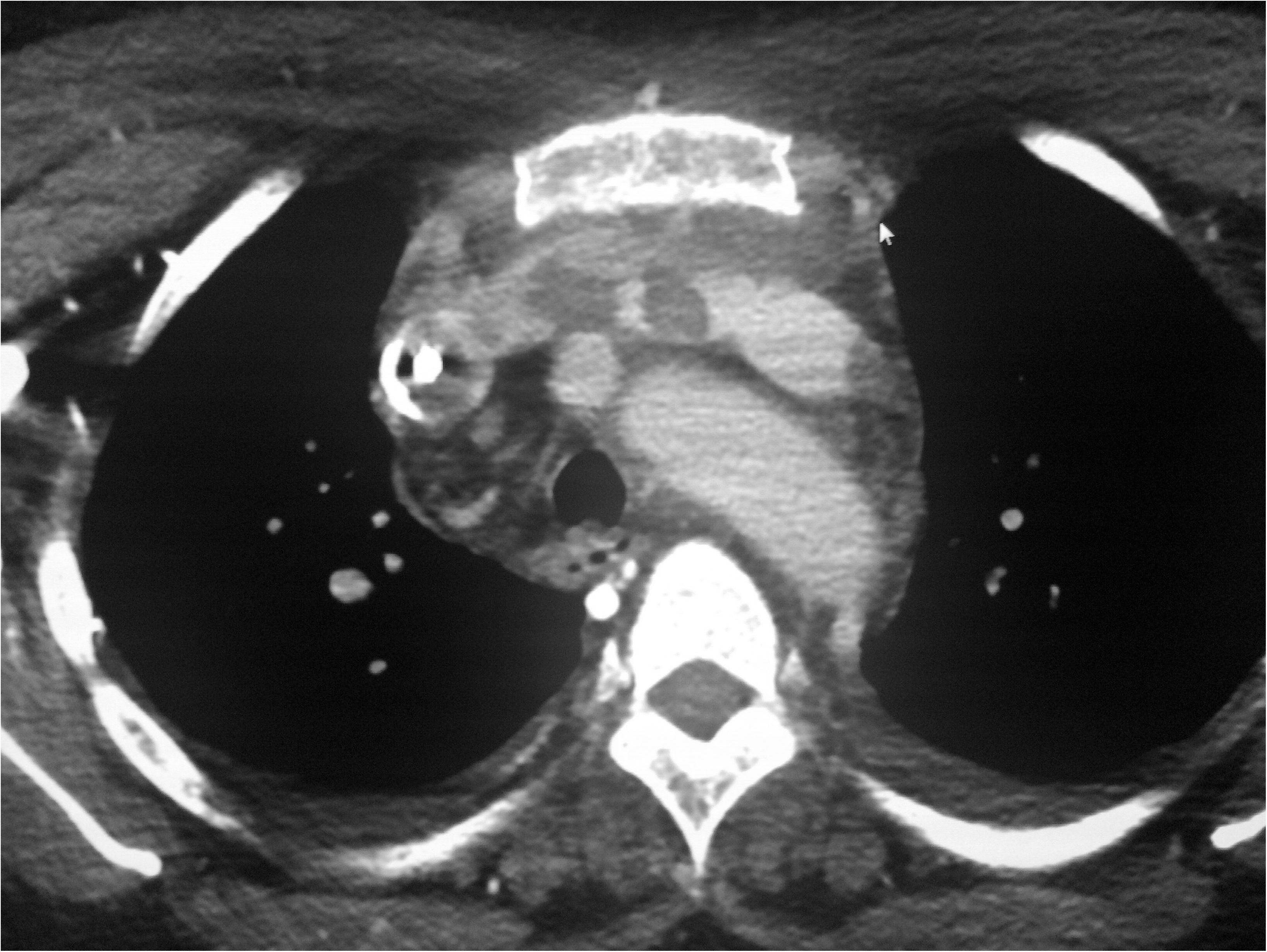
CT Venogram for preprocedural evaluation of patient with right sided port and left sided clot burden (clot within brachiocephalic pictured here).

**Figure 2:**
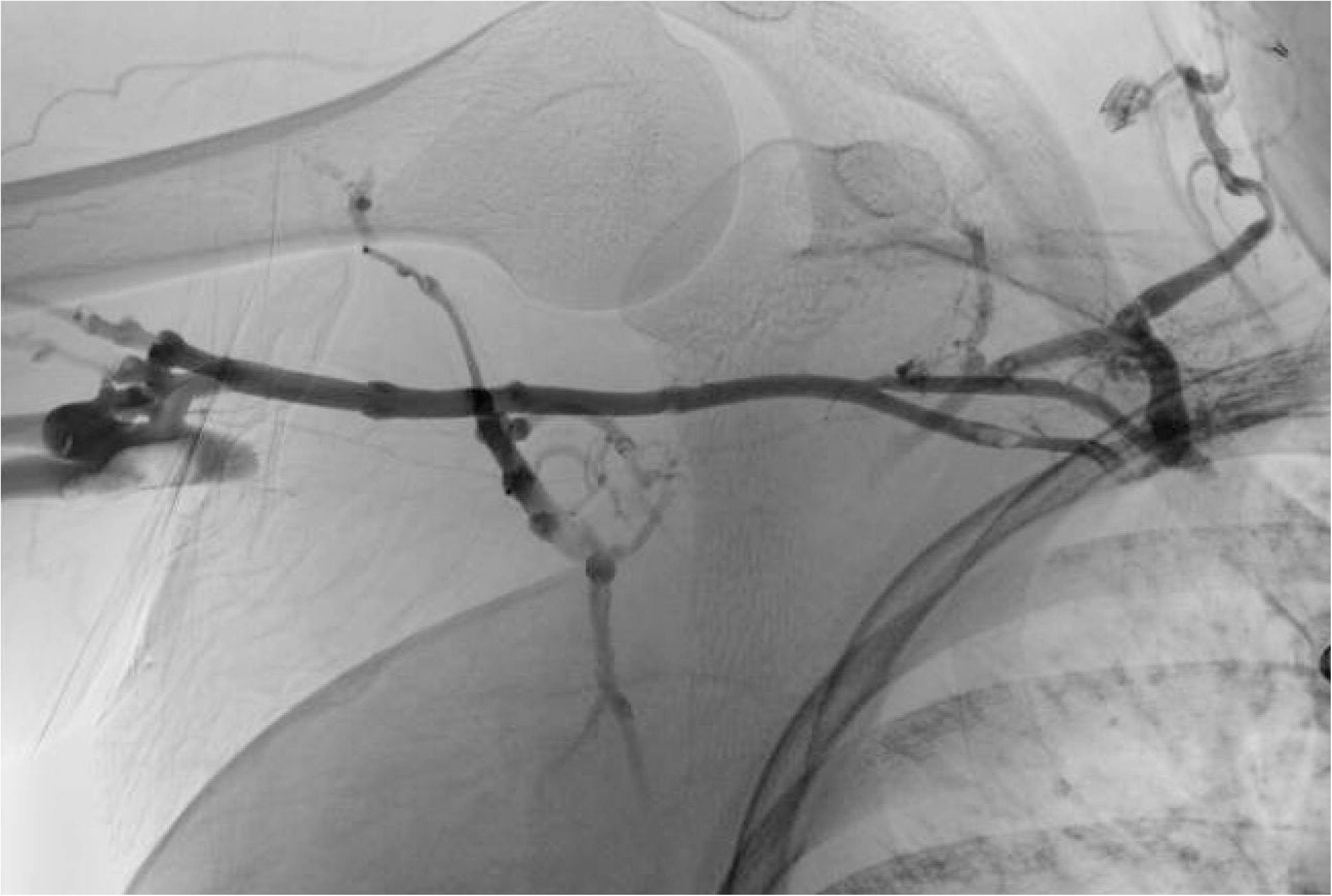
Fluoroscopic venogram images of the right upper extremity, prior to treatment, demonstrating extensive thrombosis

**Figure 3:**
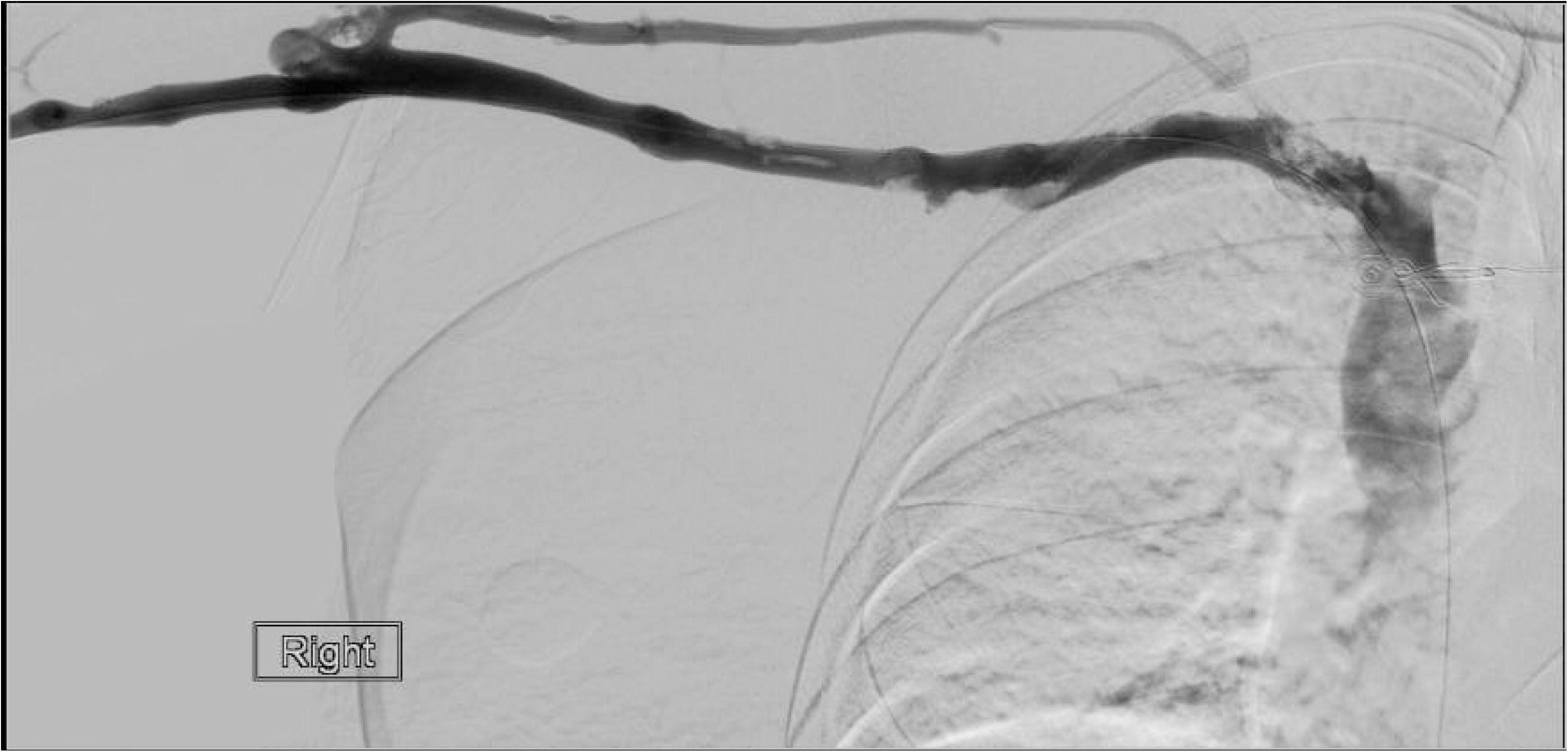
Fluoroscopic venogram images of the right upper extremity after successful recanalization protocol as described above

There were 9 total procedures performed on 7 distinct patients, with patient characteristics as noted in Table 1. Of the 9 procedures, 7 occurred in the setting of current CVC. Of the current CVCs, all were totally implantable central venous access devices (port catheters), and 5 were ipsilateral to the thrombus (71%). There were 2 procedures that occurred in the absence of a contemporaneous catheter, however both of these patients had multiple prior central venous catheters.

In follow up, one patient had a severe complication possibly related to embolization of the large thrombus burden, and expired. The family declined autopsy, leaving the cause of death speculative. One patient who did not initially have recalcitrant stenosis and therefor did not have a stent placed during the initial procedure, had recurrent stenosis and thrombosis 6 months after the initial procedure, and had repeat pharmomechanical thrombolysis performed, with stent placement at that time. The other 7 procedures (78%) were without complication, and the upper extremity and central venous systems remained patent at most recent follow-up, now up to 30 months post procedure.

## Discussion

Almost 30% of adult sickle cell disease patients have experienced central venous catheter-related thromboembolism. A few studies have identified that port catheters are associated with fewer complications in comparison with CVC^11^. However, in our small series, 6 of the 9 procedures presented with port catheter related thrombosis. Other complications of port catheters have also been demonstrated. We have noted patients in whom the port is accessed with an apheresis needle, as opposed to a non-coring needle, resulting in the puncture of the port diaphragm (Figure 4).

**Figure 4:**
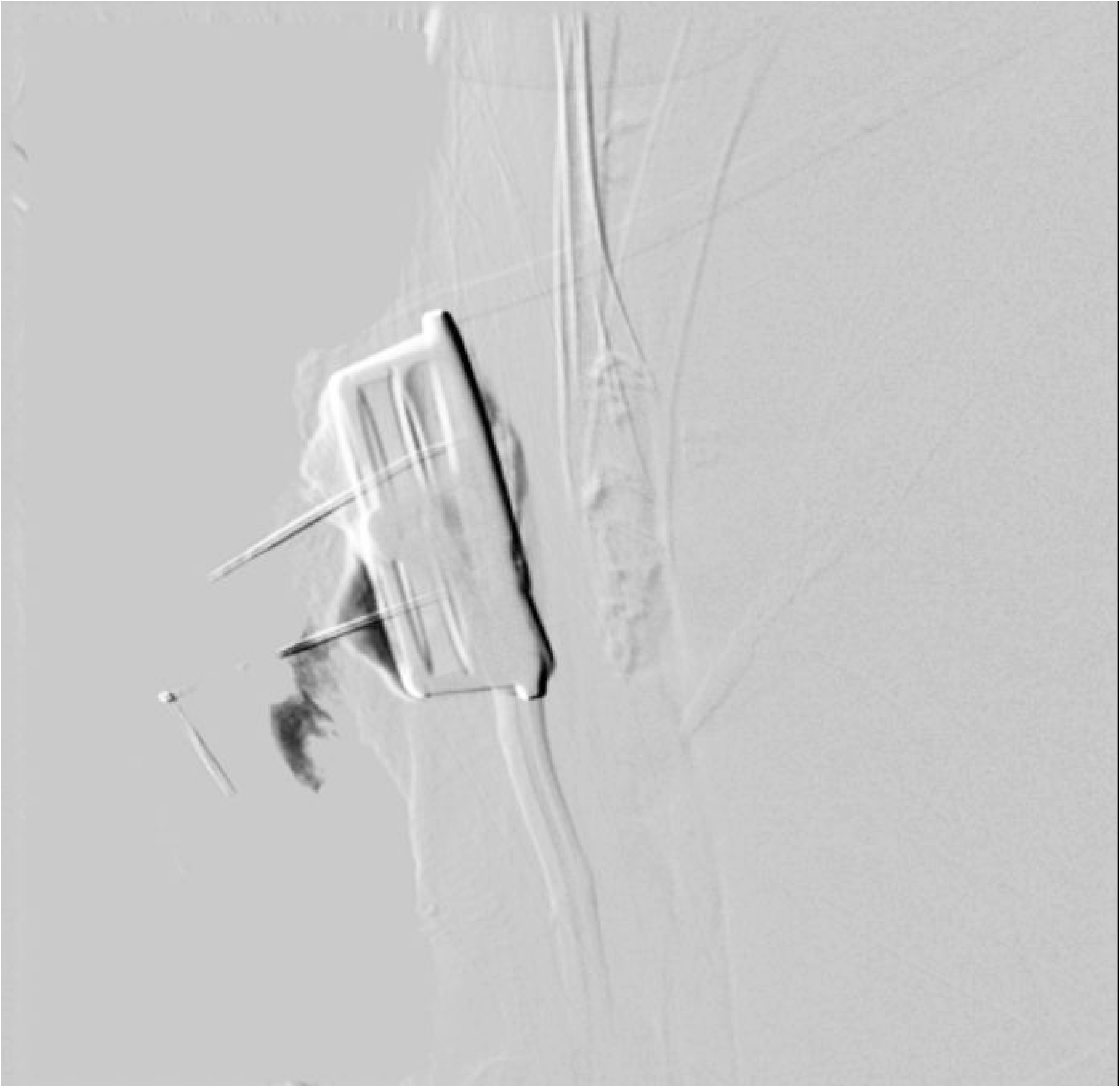
Contrast injection of a port catheter previously accessed with a pheresis needle, resulting in port well disruption and contrast extravasation

At present, the only pharmacological treatment to reduce thrombosis in the setting of SCD is hydroxyurea, which increases HbF percentage. Other treatments like analgesics, laxatives, and anxiolytics are indicated for the control of symptoms. Gene therapy is currently yielding promising results, although it’s current usage is limited by many factors including the high cost and paucity of HLA matched donors. More research is needed to improve the availability and cost-effectiveness of gene therapy.

Prior reports of treatment for catheter related thrombosis are limited. Individual case studies have demonstrated successful thromboloysis, however these have limited follow up. Given the pro thrombotic nature of sickle cell disease, in combination with the pro thrombotic nature of central venous catheters, novel solutions for pheresis access and maintenance of sickle cell disease remains an area in need of further research and innovation. For now, a multidisciplinary team should ideally be involved for the treatment of SCD.

## Conclusion

With the knowledge that CVCs are associated with increased venous complications, multidisciplinary decisions about their necessity and appropriate use can help to limit complications. In the circumstance of severe catheter related thrombosis, we present a protocol which can be considered for therapeutic use.

## Data Availability

All HIPAA compliant data is provided in the presented manuscript and tables.

